# Discordant Obesity Severity Classification Between the Edmonton Obesity Staging System and the Lancet Commission Model

**DOI:** 10.64898/2026.03.16.26348463

**Authors:** Tobias Hagemann, Arya M. Sharma, Matthias Blüher, Anne Hoffmann

## Abstract

**Objective:** BMI alone does not capture obesity-related health heterogeneity. The Edmonton Obesity Staging System (EOSS) grades obesity severity based on comorbidities and functional impairment, whereas the Lancet Commission Diagnostic Model for Obesity (DMO) distinguishes preclinical from clinical obesity based on organ dysfunction. We assessed whether both frameworks identify overlapping phenotypes and how they classify obesity severity.

**Methods:** A modified EOSS and DMO were applied to the UK Biobank (N ≈ 411,000). Stage distributions, cross-classification, and the impact of combining BMI with fat distribution on obesity categorization were analyzed.

**Results:** About one quarter of participants were classified with obesity under both frameworks. Most were assigned to advanced stages, with high concordance for established disease. Differences were most pronounced in early stages: DMO captured a broader spectrum of mild/subclinical organ dysfunction, whereas EOSS emphasized established disease with prognostic relevance. Discrepancies reflected differences in operationalization of e.g. metabolic, cardiovascular, and mental health. Obesity thresholds influenced classification, with ∼50% reclassified when BMI was combined with different fat distribution parameters, highlighting sensitivity of early-stage assignment.

**Conclusion:** EOSS and DMO provide complementary perspectives on obesity severity. Integrating EOSS’s prognostic granularity with DMO’s multidimensional approach may improve risk stratification and identify individuals most suitable for intensive interventions.

**STUDY IMPORTANCE:** *What is already known?:*

- BMI alone poorly reflects obesity-related health risk; comorbidities, organ dysfunction, and functional impairments are crucial for precise staging.
- Two major frameworks exist: EOSS focuses on prognostic severity, while DMO identifies early/preclinical obesity—but their agreement and clinical implications were unclear.

*What does this study add?:*

- Demonstrates that EOSS emphasizes established disease and prognostic severity, whereas DMO captures a broader spectrum of early or subclinical organ dysfunction, revealing distinct phenotypes within the same BMI-defined population.
- Highlights that combining BMI with anthropometric measures can reclassify up to ∼50% of individuals, illustrating the sensitivity of early-stage assignment to diagnostic thresholds.

*How might these results change the direction of research or the focus of clinical practice?:*

- Integrating EOSS’s prognostic detail with DMO’s broad, multidimensional approach enables targeted intervention, helping clinicians prioritize patients for intensive obesity management or treatment.
- Provides evidence for harmonizing obesity classification beyond BMI, emphasizing the need for multidimensional assessment in both research cohorts and routine clinical practice.

## 1 Introduction

Obesity is a chronic, complex, and relapsing multisystem disease recognized by the World Health Organization and numerous medical societies (1–4). Early diagnosis and appropriate staging are crucial to guide personalized treatment strategies and improve clinical outcomes (5, 6). Defining disease severity, however, remains challenging. Within the ICD framework, the diagnosis and grading of obesity still rely solely on body mass index (BMI) cut-off values (4). BMI-based classification may misclassify adiposity and does not adequately capture organ dysfunction, functional impairment, or individual health risk (7).

As proposed by the European Association for the Study of Obesity (EASO) and others (1–3, 8, 9), it is recommended to assess medical, functional, and psychological impairments alongside anthropometric measures to better identify individual health risks and avoid under-treatment (1, 8, 10–14). The Edmonton Obesity Staging System (EOSS), introduced in 2009 (15), exemplifies this approach by proposing a simple clinical and functional staging system. EOSS describes the morbidity and functional limitations associated with excess weight on a five-point ordinal scale, where stage 0 indicates no apparent risk factors; stage 1 reflects subclinical risk factors; stage 2 involves established chronic diseases; stage 3 signifies end-organ damage; and stage 4 denotes severe disabilities resulting from obesity-related conditions. Importantly, EOSS has been associated with increased cardiovascular and all-cause mortality (14).

In 2025, the Lancet Commission on Clinical Obesity proposed a new Diagnostic Model for Obesity (DMO), emphasizing the impact of excess adiposity on organ and tissue function (12). The DMO recommends using BMI (≥ 30 kg/m²) only for initial screening. Comprehensive assessment of obesity should confirm excess fat accumulation should by either direct measurement of body fat or by at least one anthropometric criterion, including waist circumference (WC), waist-to-hip ratio (WHR), or waist-to-height ratio (WHtR). Only in individuals with very high BMI (>40 kg/m^2^) can excess adiposity be pragmatically assumed, and further confirmation is not required. The DMO further differentiates between preclinical and clinical obesity. Preclinical obesity is characterized by excessive fat accumulation without current organ dysfunction or limitations in daily activities but with an increased risk for obesity-related diseases. In contrast, clinical obesity involves measurable organ or tissue dysfunction and/or significant age-adjusted restrictions in daily functioning resulting from excess adiposity. The primary aim of this study was to perform a cross-classification of individuals with obesity using DMO and EOSS, to identify how these systems distinguish those at highest risk for complications and premature mortality (e.g., clinical obesity in the DMO framework) from those who, despite excess adiposity, maintain preserved health (preclinical obesity). Building on the DMO framework, which requires confirmation of excess adiposity beyond BMI alone, we hypothesized that combing BMI with additional anthropometric measures would result in a reclassification across staging systems. We therefore investigated how various anthropometric parameters affect obesity categorization. To accomplish this, we utilized data from the UK Biobank (UKBB) (16), a large, well-characterized prospective cohort comprising approximately 500,000 participants. This extensive dataset enables a comprehensive comparison of classification systems and their potential implications for clinical practice.

## 2 Methods

### 2.1 Cohort Description

The UKBB cohort consists of a total of 500,000 participants, as described elsewhere (16). Only participants self-identifying as “British”, “Irish”, or “any other White background” were included in the analysis. Participants of other ethnicities (e.g., “Asian or Asian British”, “Black or Black British”, “Mixed”, or “Chinese”) were excluded due to small sample sizes and to further minimize potential ethnic bias. Individuals with diagnoses such as cancer (UKBB field ID 2453), alcoholic liver disease (UKBB field ID 131658), alcoholism (International Statistical Classification of Diseases and Related Health Problems (ICD)-10: F10), drug abuse (ICD-10: F19), pregnancy at the time of data collection (UKBB field ID 3140), and toxic liver disease (UKBB field ID 131660) were excluded. This approach resulted in a cohort of N = 411,445 participants (54% women, age 56.5 ± 8 years). According to the WHO BMI classification (17), participants were categorized as underweight (N = 1,933), normal weight (N = 134,884), overweight (N = 175,792), and obese (N = 98,836). A detailed description of the cohort is provided in Suppl. Table 1.

### 2.2 Classification Based on the modified Edmonton Obesity Staging System

The criteria for assigning EOSS stages were adopted from the Clinical Obesity Chronic Disease Dashboard for obesity (18), a modified version from the original EOSS framework (15) providing detailed operational definitions of obesity-related complications rather than the general concept described in the original framework. Obesity was defined as BMI ≥ 30 kg/m² according to WHO criteria. Clinical parameters, including cutoffs, ICD codes, and UKBB field numbers used for EOSS implementation are provided in Suppl. Table 2; concomitant medications are listed in Suppl. Table 3. ICD-9 codes provided by Clinical Obesity Chronic Disease Dashboard were translated into ICD-10, as documented in Suppl. Table 4. Hypertension and diabetes were defined according to the Canadian Prime Care Sentinal Surveillance Network (CPCSSN) (19), with HbA1c >7% used as the laboratory criterion for diabetes due to unavailable fasting glucose data in UKBB. CPCSSN hypertension was set to missing, if any of the data matched any of the CPCSSN exclusion criteria. The estimated glomerular filtration rate (eGFR) was computed using the CKD-EPI creatinine equation (20). Although the original EOSS framework considers both mental health and functional limitations for staging, the modified EOSS (18) did not include these. In our analysis, we incorporated activities of daily living (ADL), by combining self-reported mobility and daily activity limitations into a single ADL category, and mental health, assessed using the General Anxiety Disorder-7 (GAD-7) (21) and Patient Health Questionnaire-9 (PHQ-9) (22) scores, into the EOSS classification. Each diagnostic criteria generated a score (N = 10), and the highest score determined the final EOSS stage. Participants with < 5 available scores were excluded, mainly due to missing kidney, ADL and mental health scores.

### 2.3 Classification Based on the Lancet Commissiońs Diagnostic Model for Obesity

Obesity was defined as BMI ≤ 30 kg/m² and body fat percentage >25% for males or >30% for females, following the Lancet Commission (12). Participants were classified as preclinical or clinical obesity based on evidence of organ or tissue dysfunction, including signs, symptoms, or diagnostic test results. Clinical obesity was assigned when any diagnostic criterion was met. Diagnostic criteria, including parameter thresholds, comorbidities, ICD codes, and UKBB field numbers, are provided in Suppl. Table 5. ADL were included in the DMO classification using the same self-reported measures as in EOSS. Each organ system was scored 1 if any diagnostic criterion was met, 0 otherwise; participants with any score of 1 were classified as clinically obese.

### 2.4 Statistical Analyses

Dates are presented as mean ± SD. Correlations between clinical obesity-related parameters were assessed using univariate Spearman correlation analyses. Multiple testing corrections were applied by controlling the false discovery rate (FDR) appropriate for the sample size. UKBB data analysis was performed in R (v4.4.0) using the Research Analysis Platform (RAP), which is enabled by DNAnexus technology and powered by Amazon Web Services (AWS).

## 3 Results

### 3.1 Classification of Obesity Grades according to the Edmonton Obesity Staging System

To investigate the distribution of obesity severity the UKBB cohort, participants were categorized according to the modified EOSS. Overall, 24% of UKBB probands were classified as having obesity (BMI ≥ 30 kg/m^2^) (Figure 1A). When further stratifying individuals with obesity (Figure 1B), only 0.4% fell into EOSS stage 0, indicating no apparent health impairments despite obesity. An additional 8.4% exhibited obesity-related subclinical risk factors (stage 1). Most participants with obesity were assigned to more advanced stages: 42.3% to stage 2 with established obesity-related chronic diseases and 48.9% to stage 3 with end-organ damage. EOSS stage 4 could not be assigned because UKBB lacks data on end-stage disabilities typically observed in clinical settings rather than in population-based cohorts.

**Figure 1:**
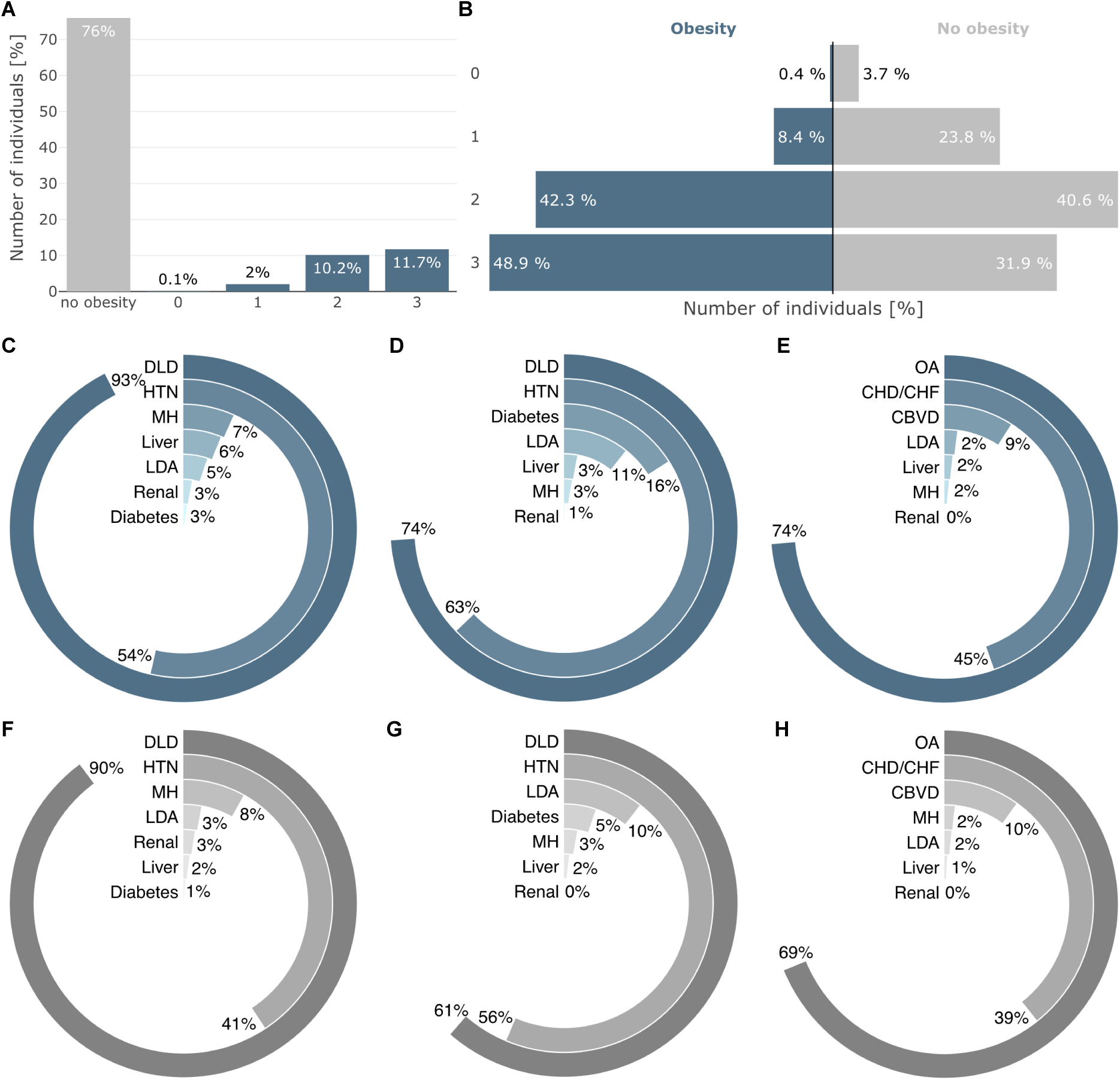
Edmonton Obesity Staging System and diagnostic criteria distribution. (A) Classification of the UK Biobank (UKBB) cohort according to the Edmonton Obesity Staging System (EOSS). The number of individuals is presented as percentages for those without obesity (BMI < 30 kg/m^2^; gray), as well as for individuals with obesity (blue), differentiated into the EOSS stages 0-3. (B) Percentage distribution of the four EOSS stages among individuals with and without obesity. (C-E) Percentage distribution of the occurrence of comorbidities contributing to the classification of patients with obesity into EOSS (C) stage 1, (D) stage 2, and (E) stage 3. (F-H) Percentage distribution of diagnostic criteria in individuals without obesity, classified into (F) stages 1, (G) stage 2, and (H) stage 3. (C-H) Only comorbidities that defined stage allocation are shown; comorbidities with non-determining scores are not included. Percentages indicate their relative contribution. CBVD: cerebrovascular disease; CHD/CHF: coronary heart disease or congestive heart failure; DLD: dyslipidemia; HTN: hypertension; LDA: limitations of daily activities; MH: Mental health; OA: osteoarthritis.

Although originally designed for patients with obesity, we also applied EOSS to participants with BMI <30 kg/m² to evaluate differences in comorbidity burden (Figure 1B). Compared to individuals with obesity, those without obesity were more often categorized into lower modified EOSS stages (+3,3% in stage 0, +15.4% in stage 1) and less frequently into higher stages (−1.7% in stage 2, -17% in stage 3).

We further examined the prevalence of comorbidities contributing to stage assignment. Only conditions directly determining stage assignment are displayed, while a complete overview is provided in Supplementary Table 6. In participants with obesity, dyslipidemia and hypertension were the most common conditions contributing to stage 1 (93% and 54%) (Figure 1c) and stage 2 (74% and 63%) (Figure 1D). Limitations in ADL and mental health impairments were observed in 5% and 7% of individuals in stage 1, respectively, and in 11% and 3% in stage 2. Liver-related diseases were present in 6% of participants in stage 1 but only 3% in stage 2, whereas type 2 diabetes (T2D) increased from 3% to 16% in these stages. In stage 3 (Figure 1E), explained by severe diseases, osteoarthritis predominated (74%), followed by coronary artery disease or congestive heart failure (45%) and cerebrovascular disease (9%). Renal impairment remains relatively uncommon across all modified EOSS stages. Among participants without obesity, the relative order of comorbidities prevalence across stages remained largely similar, although absolute frequencies were (Figure 1F-H).

### 3.2 Severity of Obesity according to the Lancet Commissiońs Diagnostic Model for Obesity

Applying the Lancet Commissiońs DMO classification, 23.4% of participants were identified as having obesity (BMI ≥ 30 kg/m^2^, body fat > 25/30% (M/F)) (Figure 2A). Among individuals with obesity, clinical obesity occurred about 8.6 times more frequently than preclinical obesity (Figure 2B). Individuals with obesity showed a 12% higher rate of clinical obesity classification than those without obesity, indicating greater impairment of organ or tissue function and ADL (Figure 2B). The most frequent conditions among individuals with clinical obesity (Figure 2C) were metabolic (73%), cardiovascular (58%), and musculoskeletal dysfunction (51%). Other contributors included respiratory (31%) or renal dysfunction (28%), and LDA (17%). In individuals without obesity (Figure 2D), these criteria occurred at substantially lower frequencies, particularly for cardiovascular (−29%), respiratory (−26%), metabolic (−18%), musculoskeletal (−17%), renal impairments (−10%), and LDA (−6%). Dysfunction of the liver, central nervous system, urinary, lymphatic, or reproductive systems were rare or absent in both groups. A detailed overview of diagnostic criteria and phenotypic characteristics for each classification is provided in Suppl. Table 7.

**Figure 2:**
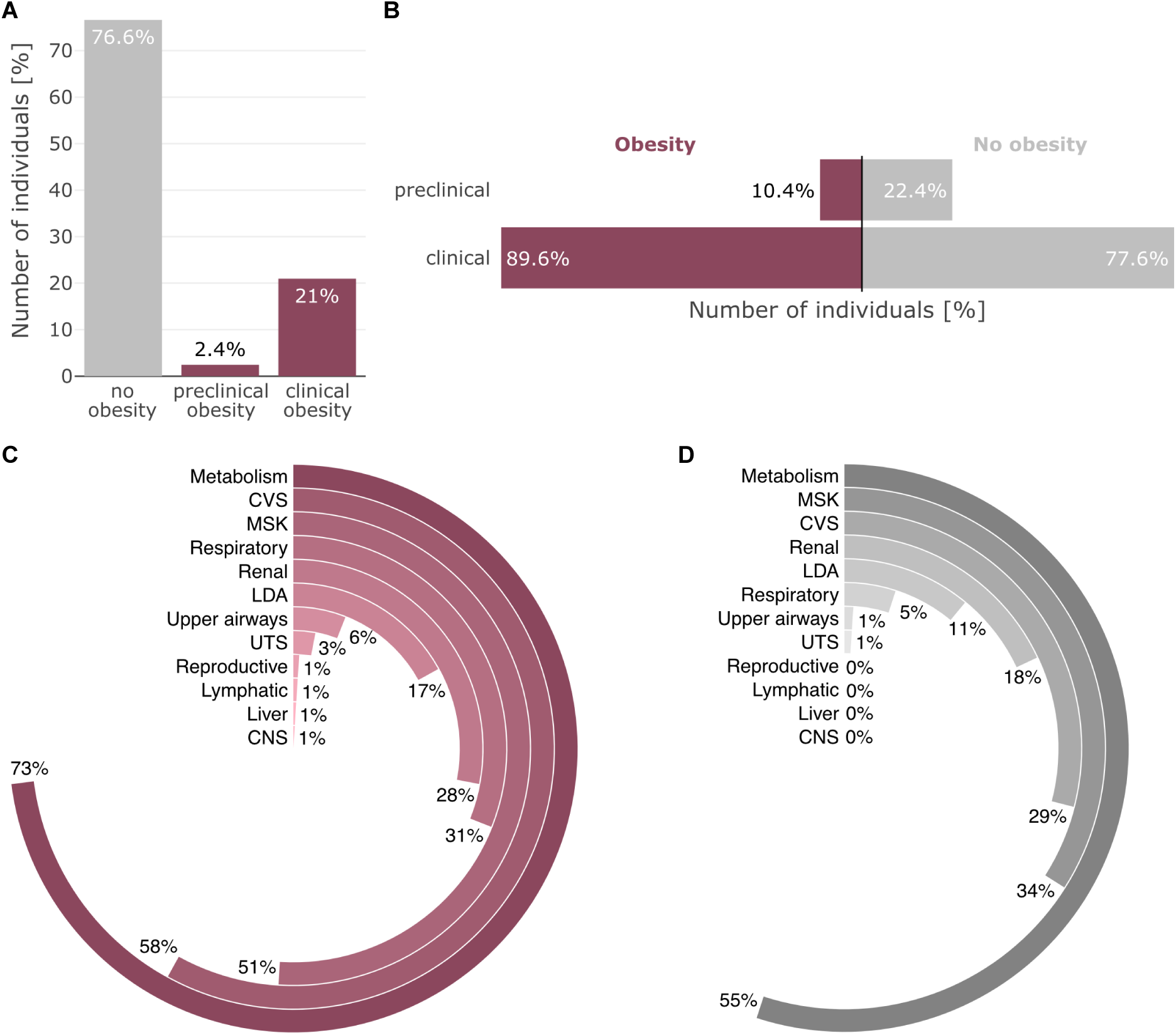
Diagnostic Model for Obesity classification and diagnostic criteria distribution. (A) Classification of the UKBB cohort according to the Diagnostic Model for Obesity (DMO). The number of individuals is presented as percentages for those without obesity (BMI < 30 kg/m^2^ and body fat ≤ 25% in males; ≤ 30% in females; gray), as well as for individuals with obesity (red), differentiated into preclinical and clinical obesity. (B) Percentage distribution of preclinical and clinical classifications among individuals with and without obesity. (C) Percentage distribution of diagnostic criteria contributing to the classification of patients with obesity as clinically obese. (D) Percentage distribution of diagnostic criteria in individuals without obesity. (C-D) Percentages indicate their relative contribution. CNS: central nervous system; CVS: cardiovascular system; LDA: limitations of daily activities; MSK: musculoskeletal; UTS: urinary tract system.

### 3.3 Differences Between the Classification Systems in Defining Obesity Severity

To compare how participants are classified by the modified EOSS and DMO, Figure 3A presents the absolute numbers of participants assigned to each category. Due to the different criteria for defining obesity (EOSS: BMI ≥ 30 kg/m^2^; DMO: BMI ≥ 30 kg/m^2^ and body fat > 25/30% (M/F)), some individuals classified as having no obesity by DMO were categorized into EOSS stages 0-3. Notably, a substantial proportion of these individuals were assigned to stage 2 (N = 1,154) and stage 3 (N = 1,071). Although many participants fall into comparable severity categories (e.g., EOSS stages 1-3 and clinical obesity), differences remain. Of the individuals classified as preclinical obesity by DMO (N = 10,005), only 279 were assigned to EOSS stage 1. In contrast, most participants in EOSS stage 0 (279 of 420) were classified as preclinical obesity by DMO, indicating that EOSS assigns fewer individuals to the lowest severity category.

**Figure 3:**
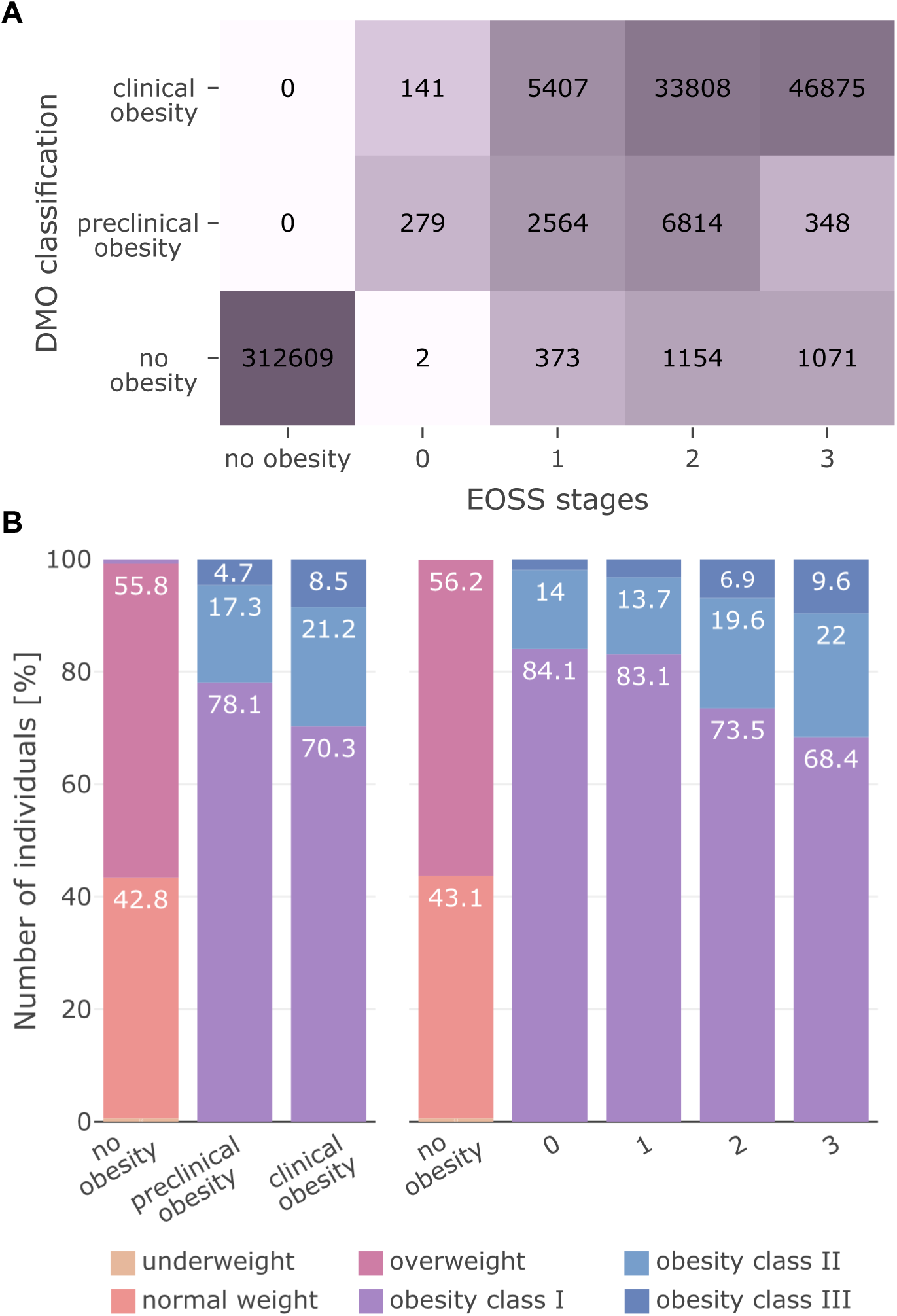
Patient and BMI classes distribution across the diagnostic classification systems. (A) Heatmap displaying the absolute number of patients in the UKBB, with a comparative overview of patient counts across EOSS and DMO classification groups. (B) Percentage distribution of World Health Organization-defined BMI classes within each EOSS and DMO classification group.

To evaluate how WHO BMI categories reflect obesity-related health status, we examined their distribution across DMO and EOSS classifications (Figure 3B). Higher obesity classes were associated with a greater prevalence of advanced modified EOSS stages and clinical obesity in DMO. However, a considerable proportion of individuals with obesity class II or III remaining categorized as preclinical obesity (DMO) or remained in the lower EOSS stages (0–1).

Additionally, we examined obesity-related clinical parameters not utilized for classification across obesity severity levels (Suppl. Figures 1, 2). Obesity severity defined by DMO and EOSS was more closely associated with age and MRI-assessed visceral adipose tissue (VAT) volume than with BMI or other anthropometric measures. Age increased with higher obesity stages but showed little association with fat distribution parameters, suggesting that disease burden rises with age independently of overall fat mass. Although MRI-assessed VAT levels strongly correlate with BMI and other fat distribution measures, it showed a much clearer stepwise increase across obesity severity stages. In contrast, body weight, WC, WHR, or WHtR, and MRI-assessed subcutaneous adipose tissue (SAT) volumes remained relatively stable.

### 3.4 Effect of Adding Anthropometric Criteria to Categorize Obesity Severity

The choice of clinical parameters and cutoff values for defining obesity remains debated. Recent recommendations, including those from the Lancet Commission on Obesity and EASO, suggested incorporating measures of excess adiposity beyond BMI (1, 12). Specifically, the Lancet Commission suggests incorporating either assessments of body fat or at least one anthropometric criterion such as WC, WHR, or WHtR, in addition to BMI. For individuals with BMI ≥40 kg/m², they proposed that excess adiposity may be assumed pragmatically.

Accordingly, we evaluated several proposed parameters and cutoff thresholds for their effect on classification outcomes. Parameters tested included (I) BMI ≥ 30 kg/m² alone (in line with the WHO (23)), or the Lancet Commission recommendations (12): BMI ≥ 40 kg/m², or BMI ≥ 30 kg/m² in combination with (II) body fat > 25/30% (M/F), (III) WC > 1021/88 cm (M/F), (IV) WHR > 0.9/0.85 (M/F), or (V) WHtR >0.5. Furthermore, as recommended by the EOSS and others (1, 11, 14) a lower (VI) BMI threshold of ≥ 25 kg/m² was also tested, either alone or in combination with the same additional parameters suggested by the Lancet Commission: (VII) body fat, (VIII) WC, (IX) WHR, or (X) WHtR. The European guidelines for obesity management (10) additionally proposed using (XI) a WC thresholds of ≥ 94/80 cm (M/F) alone or/and (XII/XIII) combined with BMI ≥ 25 kg/m^2^. Finally, (XIV) BMI ≥ 30 kg/m², or BMI ≥ 25 kg/m² in combination with a WHtR > 0.5 was tested following the guidance of the EASO (1).

As shown in Figure 4, the proportion of participants classified with obesity varied substantially depending on the chosen parameter combinations and thresholds, with consistent patterns across both diagnostic systems. Stricter criteria, particularly BMI ≥ 30 kg/m² combined with (II) body fat, (III) WC or (V) WHtR, produced largely stable classifications comparable with (I) BMI ≥ 30 kg/m² alone. In contrast, combining BMI with (IV) WHR was the least restrictive approach, leading to the largest increase in individuals classified with obesity.

**Figure 4:**
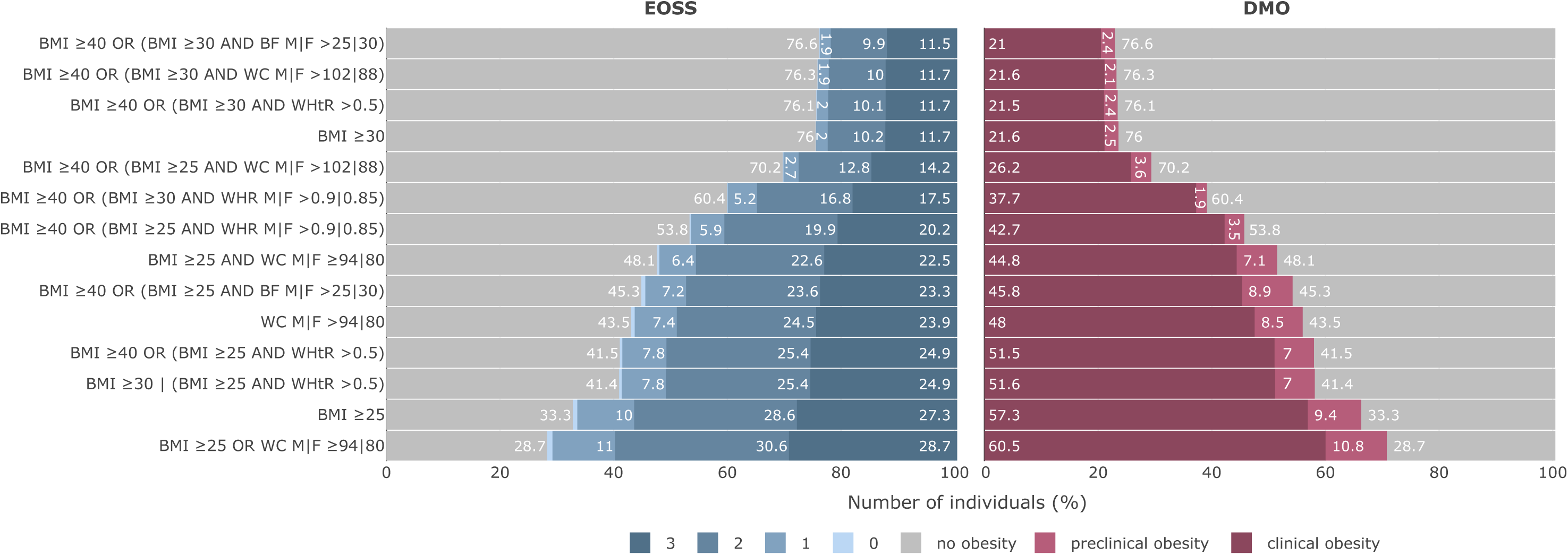
Impact of clinical parameter selection on diagnostic classification systems. Assessment of the impact of different clinical parameter cutoffs on the classification of UKBB participants into no obesity and obesity groups for EOSS (left) and DMO (right). Different cutoffs and their combinations were evaluated for body mass index (BMI, kg/m²), waist circumference (WC, cm), waist-to-height ratio (WHtR), and waist-to-hip ratio (cm), with certain cutoffs being gender-specific, adjusted separately for males (M) and females (F). Values are shown only if >1%.

The highest prevalence of obesity was observed using (VIII) BMI ≥ 25 kg/m^2^ or WC ≥ 94/80 cm (M/F), followed by (VI) BMI ≥ 25 kg/m^2^ alone. Under the BMI ≥25 kg/m² threshold, additional parameters influenced classifications more strongly, with WC and WHR yielded comparatively lenient results, while body fat and WHtR reduced obesity prevalence more substantially.

## 4 Discussion

Obesity is a complex and chronic condition that remains substantially underdiagnosed despite its well-documented health risks. In the UK Biobank, only 9.8% of participants had a recorded ICD-10 diagnosis of obesity, underscoring the gap between clinical documentation and the actual burden of adiposity. In this study, we therefore performed a cross-classification of obesity severity using both modified EOSS and the DMO framework to evaluate how these systems capture adiposity-related disease burden.

Approximately one quarter of participants were classified as having obesity under both frameworks. However, most individuals with obesity were assigned to advanced stages of disease burden, with nearly half of participants reaching EOSS stage 3 and clinical obesity predominating within the DMO classification. Similar distributions have been reported in other cohorts, although differences between the UKBB, the Northern Alberta Primary Care Research Network and the All of Us cohort (24) likely reflect variation in age structure, recruitment strategies, and underlying health status. Nevertheless, the consistently high proportion of individuals with advanced disease stages highlights the substantial health burden associated with excess adiposity.

When applying both classification systems to individuals with and without obesity, participants with adiposity consistently showed a markedly higher prevalence of advanced disease stages, whereas individuals without obesity were predominantly assigned to lower stages. These findings emphasize the strong association between excess adiposity and multi-system disease burden.

Both frameworks identify metabolic and cardiovascular impairments as the most prevalent domains among individuals with obesity, consistent with previous reports (18, 25). They differ, however, in operationalization. The modified EOSS uses defined clinical thresholds and established end-organ damage, including diabetes, hypertension, dyslipidemia, and cardiovascular disease. In contrast, the DMO captures broader metabolic dysfunction, such as hyperglycemia and abnormal lipid profiles, without using diabetes as a direct staging determinant. DMO also includes musculoskeletal, respiratory, upper airway, CNS, urinary, and lymphatic dysfunction, representing a broader morbidity spectrum. In EOSS, musculoskeletal disease mainly reflects osteoarthritis in stage 3, while mental health parameters are uniquely incorporated in EOSS.

Renal impairment further illustrates differences in the detection of early abnormalities between the two systems. In the DMO framework, renal abnormalities were present in more than a quarter of individuals with clinical obesity, whereas ≤3% of participants met renal criteria in the modified EOSS. This discrepancy likely reflects the broader inclusion of mild or early renal abnormalities in DMO, whereas the modified EOSS relies on stricter thresholds based on eGFR and albuminuria, thereby capturing more advanced disease. Liver disease was identified at low frequencies in both frameworks, likely reflecting substantial underdiagnosis in population cohorts. The modified EOSS identifies hepatic involvement mainly through abnormal liver enzymes or diagnosed liver disease, while DMO includes metabolic dysfunction-associated steatotic liver disease and hepatic fibrosis as diagnostic entities. Because these conditions frequently require imaging or biopsy for confirmation, they are rarely detected in routine clinical practice (26).

Taken together, the modified EOSS provides a structured staging system emphasizing clinically established disease and organ damage, whereas the DMO framework adopts a broader, multidimensional perspective capturing functional limitations and organ dysfunction across multiple systems. Consequently, DMO identifies a wider spectrum of obesity-related impairments, while EOSS focuses more strongly on disease severity and prognostic relevance. In our cohort, fewer participants were classified as stage 0 under EOSS than as preclinical obesity under DMO, likely reflecting the stricter metabolic and psychological criteria required to meet the EOSS stage 0 definition. Cross-classification further revealed that a subset of individuals classified as non-obese in DMO were assigned to advanced EOSS stages, illustrating the conceptual differences between both frameworks. Recognizing these complementary strengths, a recent proposal suggested combining the DMO diagnostic definition with EOSS staging to integrate practical diagnosis with prognostic stratification (27).

Both frameworks share limitations. Liver disease and several organ system impairments appear underrepresented, likely reflecting underdiagnosis and limited routine screening. In addition, neither system incorporates lifestyle-related risk factors, such as dietary patterns, smoking, or alcohol consumption, despite their strong influence on obesity-related comorbidities (28). Cancer is also not included in either framework, although evidence from the UK Biobank demonstrates strong associations between both preclinical and clinical obesity in the DMO framework and multiple obesity-related cancers (29).

A foundational step for any classification system is to distinguish between individuals with and without obesity, as BMI alone does not capture fat distribution or composition. Consistent across both EOSS and DMO classifications, our analysis revealed substantial heterogeneity within WHO BMI categories: although higher obesity classes generally corresponded to more advanced disease stages, a considerable proportion of individuals with class II or III obesity were still categorized as preclinical obesity or remained in lower EOSS stages (0–1). These findings highlight that individuals with similar BMI can exhibit markedly different metabolic and functional profiles.

Current guidelines therefore recommend complementing BMI with measures of abdominal fat accumulation such as WC, WHtR, or WHR (1, 8, 10, 11, 30). Our findings underline that the choice of diagnostic thresholds is not trivial, as up to ∼50% of individuals being reclassified depending on the applied definition. However, combining BMI ≥30 kg/m² with WC, body fat, or WHtR resulted in only minor deviations from BMI ≥ 30 kg/m² alone, suggesting limited additional value of these parameters once a clear obesity threshold is reached. In contrast, combining BMI ≥30 kg/m² with WHR produced the least restrictive classifications. This likely reflects that WHR depends strongly on both waist and hip circumference and may therefore capture body shape rather than visceral fat accumulation. At lower BMI thresholds (≥25 kg/m²), additional anthropometric parameters had a stronger influence on classification, which aligns with the notion that borderline categories are more sensitive to measurement choice (31, 32). Clinically, this suggests that WC or WHR may support earlier identification of individuals at risk but may also increase overdiagnosis, whereas body fat or WHtR could prioritize specificity. These support current calls for harmonization of cutoff definitions (12, 32), emphasizing the need for evidence-based standardization to avoid inconsistencies in obesity classification across clinical and research settings.

Finally, obesity severity in our cohort was more strongly associated with age and MRI- derived VAT than with BMI or other standard anthropometric measures. While age contributed to increasing disease burden independently of fat distribution, VAT showed a clear stepwise increase across severity stages, highlighting the potential value of imaging-derived measures to complement conventional anthropometric criteria in obesity classification.

### 4.1 Limitations

A limitation of this study is that the UK Biobank does not allow assessment of whether comorbidities are directly attributable to adiposity. We therefore assumed that all documented conditions were obesity-related, which may overestimate associations between adiposity and comorbidities. This is in line with the EOSS approach that causality of a disease or condition to obesity is irrelevant (and frequently difficult to establish) for the stage categorization. In contrast, DMO places greater emphasis on obesity-related causality, which may not adequately reflect the individual obesity-related disease burden.

Another limitation concerns the implementation of the DMO classification, as its diagnostic criteria are less clearly standardized than those of the modified EOSS. Consequently, some criteria required interpretation during operationalization. In addition, we applied a modified EOSS framework based on the Clinical Obesity Chronic Disease Dashboard (18) rather than the original EOSS (15), as detailed operational definitions were required for cohort-based analyses; similar adaptations have been applied in previous studies due to data limitations (14, 18, 33). The application of both frameworks was further constrained by the availability of clinical and laboratory data in the UKBB. In addition, functional limitations were assessed using self-reported measures of daily activity and mobility, which may introduce reporting bias. Finally, the UKBB cohort is not fully representative of the general population and tends to include healthier volunteers, which may limit generalizability.

## 5 Conclusions

Our comparison of the modified EOSS and the DMO frameworks highlights their complementary roles in characterizing obesity-related disease burden. While EOSS provides structured prognostic staging based on clinically established organ damage, DMO captures a broader spectrum of functional limitations and early organ dysfunction across multiple systems. The observed discordance between both frameworks and the substantial heterogeneity within BMI categories further emphasize that BMI alone does not adequately reflect obesity severity. In addition, our findings underline the influence of diagnostic thresholds and fat distribution measures on obesity classification. Integrating the prognostic granularity of EOSS with the multidimensional perspective of DMO, together with harmonized diagnostic criteria, may improve risk stratification and support more targeted clinical management of obesity.

## Data Availability

The data used in this study are available from the UKBB (https://www.ukbiobank.ac.uk/) upon application. The analyses were performed using the UKBB dataset as available in 2025. Due to UKBB policies, the data cannot be publicly shared by the authors.

## Acknowledgements

This research has been conducted using the UK Biobank Resource under application number: 260619.

## Suppl. Table Legends

Suppl. Table 1: Detailed cohort description. Phenotypic description of the UK Biobank (UKBB) cohort analyzed, subdivided into the individual World Health Organization (WHO) BMI categories, as well as total population.

Suppl. Table 2: Revised Edmonton Obesity Staging System scoring definitions. Definitions applied for Edmonton Obesity Staging System (EOSS) staging, including clinical parameters, ICD codes, and UKBB field numbers for the respective comorbidities, adapted from Swaleh et al. (2021) (18). “n/a” denotes stages not defined for a given comorbidity.

Suppl. Table 3: EOSS scoring concomitant medications. Medications considered for revised EOSS staging, including statins, diabetes, and antihypertensive drugs.

Suppl. Table 4: International Classification of Diseases codes applied for the revised EOSS scoring. International Classification of Diseases (ICD)-9 codes as proposed Swaleh et al. (2021) (18), and their corresponding ICD-10 codes as recorded in the UKBB for the respective diseases. All ICD-10 codes are extracted from the UKBB filed ID 41270.

Suppl. Table 5: Model for Obesity classification definitions. Definitions applied for the Diagnostic Model for Obesity (DMO) classification, including clinical parameters, ICD codes, and UKBB field numbers for the respective diagnostic criteria. All ICD-10 codes are extracted from the UKBB filed ID 41270.

Suppl. Table 6: Participant characteristics across EOSS stages. Phenotypic characteristics and comorbidities of individuals stratified by revised EOSS stage.

Suppl. Table 7: Participant characteristics across DMO classes. Phenotypic characteristics and diagnostic criteria of individuals stratified by DMO categories.

## References

1. Busetto L, Dicker D, Frühbeck G, et al. A new framework for the diagnosis, staging and management of obesity in adults. Nat Med 2024;30:2395–2399.

2. Mechanick JI, Garber AJ, Handelsman Y, et al. American Association of Clinical Endocrinologists’ Position Statement on Obesity and Obesity Medicine. Endocr Pract 2012;18:642–648.

3. Bray GA, Kim KK, Wilding JPH, on behalf of the World Obesity Federation. Obesity: a chronic relapsing progressive disease process. A position statement of the World Obesity Federation. Obes Rev 2017;18:715–723.

4. World Health Organization 2024. International Classification of Diseases ICD-11 for mortality and morbidity statistics: 5B81 obesity, Accessed August 11, 2025. https://icd.who.int/browse/2024-01/mms/en#149403041. 2024.

5. Blüher M. Metabolically Healthy Obesity. Endocr Rev 2020;41:bnaa004.

6. Kopelman PG. Obesity as a medical problem. Nature 2000;404:635–643.

7. Wu Y, Li D, Vermund SH. Advantages and Limitations of the Body Mass Index (BMI) to Assess Adult Obesity. Int J Environ Res Public Health 2024;21:757.

8. Wharton S, Lau DCW, Vallis M, et al. Obesity in adults: a clinical practice guideline. Can Med Assoc J 2020;192:E875–E891.

9. American Society for Metabolic and Bariatric Surgery. Consensus Statement on Obesity as a Disease, Accessed August 11, 2025. https://asmbs.org/resources/consensus-statement-on-obesity-as-a-disease/. 2022.

10. Yumuk V, Tsigos C, Fried M, et al. European Guidelines for Obesity Management in Adults. Obes Facts 2015;8:402–424.

11. Garvey WT, Mechanick JI, Brett EM, et al. American Association of Clinical Endocrinologists and American College of Endocrinology Comprehensive Clinical Practice Guidelines For Medical Care of Patients with Obesity. Endocr Pract 2016;22:1–203.

12. Rubino F, Cummings DE, Eckel RH, et al. Definition and diagnostic criteria of clinical obesity. Lancet Diabetes Endocrinol 2025;13:221–262.

13. Palacios S, Orozco R, Garcia-Almeida JM. Beyond body mass index: redefining the diagnosis of obesity. Gynecol Endocrinol 2025;41.

14. Padwal RS, Pajewski NM, Allison DB, Sharma AM. Using the Edmonton obesity staging system to predict mortality in a population-representative cohort of people with overweight and obesity. Can Med Assoc J 2011;183:E1059–E1066.

15. Sharma AM, Kushner RF. A proposed clinical staging system for obesity. Int J Obes 2009;33:289–295.

16. Bycroft C, Freeman C, Petkova D, et al. The UK Biobank resource with deep phenotyping and genomic data. Nature 2018;562:203–209.

17. World Health Organization 2024. Obesity and overweight, Accessed August 11, 2025. https://www.who.int/en/news-room/fact-sheets/detail/obesity-and-overweight. 2024.

18. Swaleh R, McGuckin T, Myroniuk TW, et al. Using the Edmonton Obesity Staging System in the real world: a feasibility study based on cross-sectional data. CMAJ Open 2021;9:E1141–E1148.

19. CPCSSN Team. Case Definitions: Canadian Primary Care Sentinel Surveillance Network (CPCSSN). 2023;Version 2022-Q4.

20. Levey AS, Stevens LA, Schmid CH, et al. A New Equation to Estimate Glomerular Filtration Rate. Ann Intern Med 2009;150:604–612.

21. Spitzer RL, Kroenke K, Williams JBW, Löwe B. A Brief Measure for Assessing Generalized Anxiety Disorder: The GAD-7. Arch Intern Med 2006;166:1092.

22. Kroenke K, Spitzer RL, Williams JBW. The PHQ-9: Validity of a brief depression severity measure. J Gen Intern Med 2001;16:606–613.

23. WHO Consultation on Obesity (1999: Geneva S, World Health Organization. Obesity : preventing and managing the global epidemic : report of a WHO consultation. Obésité Prév Prise En Charge Épidémie Mond Rapp Une Consult OMS 2000.

24. Yao Z, Dardari ZA, Razavi AC, et al. Prevalence of clinical obesity versus BMI-defined obesity among US adults: a cohort study. Lancet Diabetes Endocrinol 2025.

25. Canning KL, Brown RE, Wharton S, Sharma AM, Kuk JL. Edmonton Obesity Staging System Prevalence and Association with Weight Loss in a Publicly Funded Referral-Based Obesity Clinic. J Obes 2015;2015:619734.

26. Neale EP, Middleton J, Lambert K. Barriers and enablers to detection and management of chronic kidney disease in primary healthcare: a systematic review. BMC Nephrol 2020;21.

27. Zahid S, Peng AW, Razavi AC, Yao Z, Blumenthal RS, Blaha MJ. Center Stage: Putting Obesity Staging Systems Into the Spotlight. Prev Chronic Dis 2025;22:250222.

28. Rassy N, Van Straaten A, Carette C, Hamer M, Rives-Lange C, Czernichow S. Association of Healthy Lifestyle Factors and Obesity-Related Diseases in Adults in the UK. JAMA Netw Open 2023;6:e2314741.

29. Leitzmann MF, Stein MJ, Baurecht H, Freisling H. Excess adiposity and cancer: evaluating a preclinical-clinical obesity framework for risk stratification. eClinicalMedicine 2025;83:103247.

30. Ross R, Neeland IJ, Yamashita S, et al. Waist circumference as a vital sign in clinical practice: a Consensus Statement from the IAS and ICCR Working Group on Visceral Obesity. Nat Rev Endocrinol 2020;16:177–189.

31. Molarius A, Seidell JC, Sans S, Tuomilehto J, Kuulasmaa K. Varying Sensitivity of Waist Action Levels to Identify Subjects with Overweight or Obesity in 19 Populations of The WHO MONICA Project. J Clin Epidemiol 1999;52:1213–1224.

32. Ghesmaty Sangachin M, Cavuoto LA, Wang Y. Use of various obesity measurement and classification methods in occupational safety and health research: a systematic review of the literature. BMC Obes 2018;5:28.

33. Kuk JL, Ardern CI, Church TS, et al. Edmonton Obesity Staging System: association with weight history and mortality risk. Appl Physiol Nutr Metab 2011;36:570–576.

