## Supplementary material for "Discordant Obesity Severity Classification Between the Edmonton Obesity Staging System and the Lancet Commission Model": Suppl. Figures

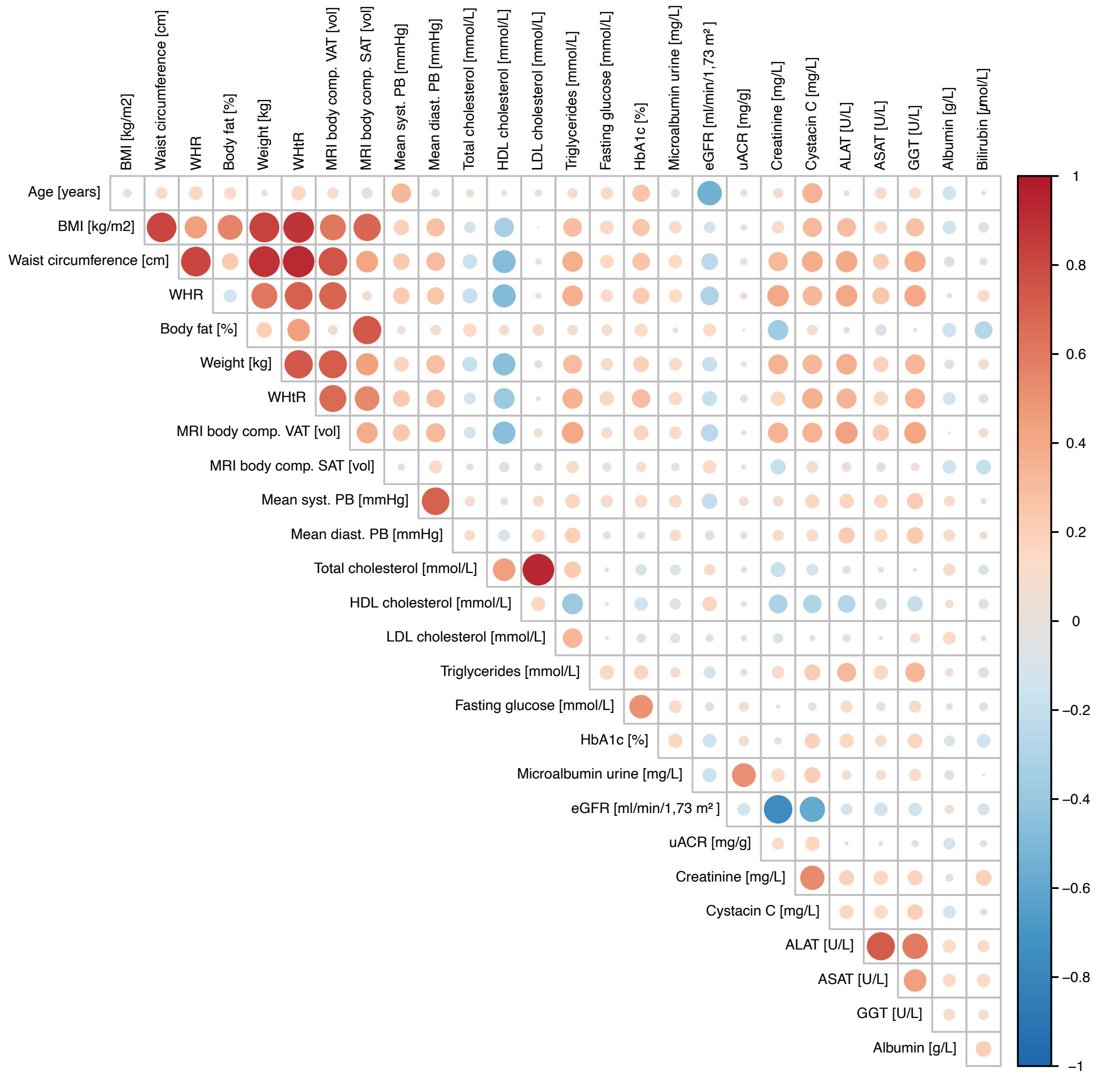

BMI [kg/m2]
Waist circumference [cm]
WHR
Body fat [%]
Weight [kg]
WHtR
MRI body comp. VAT [vol]
MRI body comp. SAT [vol]
Mean syst. PB [mmHg]
Mean diast. PB [mmHg]
Total cholesterol [mmol/L]
HDL cholesterol [mmol/L]
LDL cholesterol [mmol/L]
Triglycerides [mmol/L]
Fasting glucose [mmol/L]
HbA1c [%]
Microalbumin urine [mg/L]
eGFR [ml/min/1,73 m²]
uACR [mg/g]
Creatinine [mg/L]
Cystacin C [mg/L]
ALAT [U/L]
ASAT [U/L]
GGT [U/L]
Albumin [g/L]
Bilirubin [µmol/L]
Age [years]
1
0.8
0.6
0.4
0.2
0
−0.2
−0.4
−0.6
−0.8
−1

**Supplementary Figure 1: Correlation of clinical parameters of the UK Biobank.** Spearman correlation matrix of selected clinical parameters. The color scale ranges from red (strong positive correlation, +1) to blue (strong negative correlation, -1). Abbreviations: ALAT: alanine aminotransferase; ASAT: aspartate aminotransferase; BMI: body mass index; eGFR: estimated glomerular filtration rate; BP: blood pressure; GGT: gamma-glutamyl transferase; HbA1c: hemoglobin A1c; HDL: high-density lipoprotein; LDL: low-density lipoprotein; MRI: magnetic resonance imaging; SAT: subcutaneous adipose tissue; uACR: urinary albumin-to-creatinine ratio; VAT: visceral adipose tissue; WHR: waist-hip ratio; WHtR: waist-to-heigh ratio.

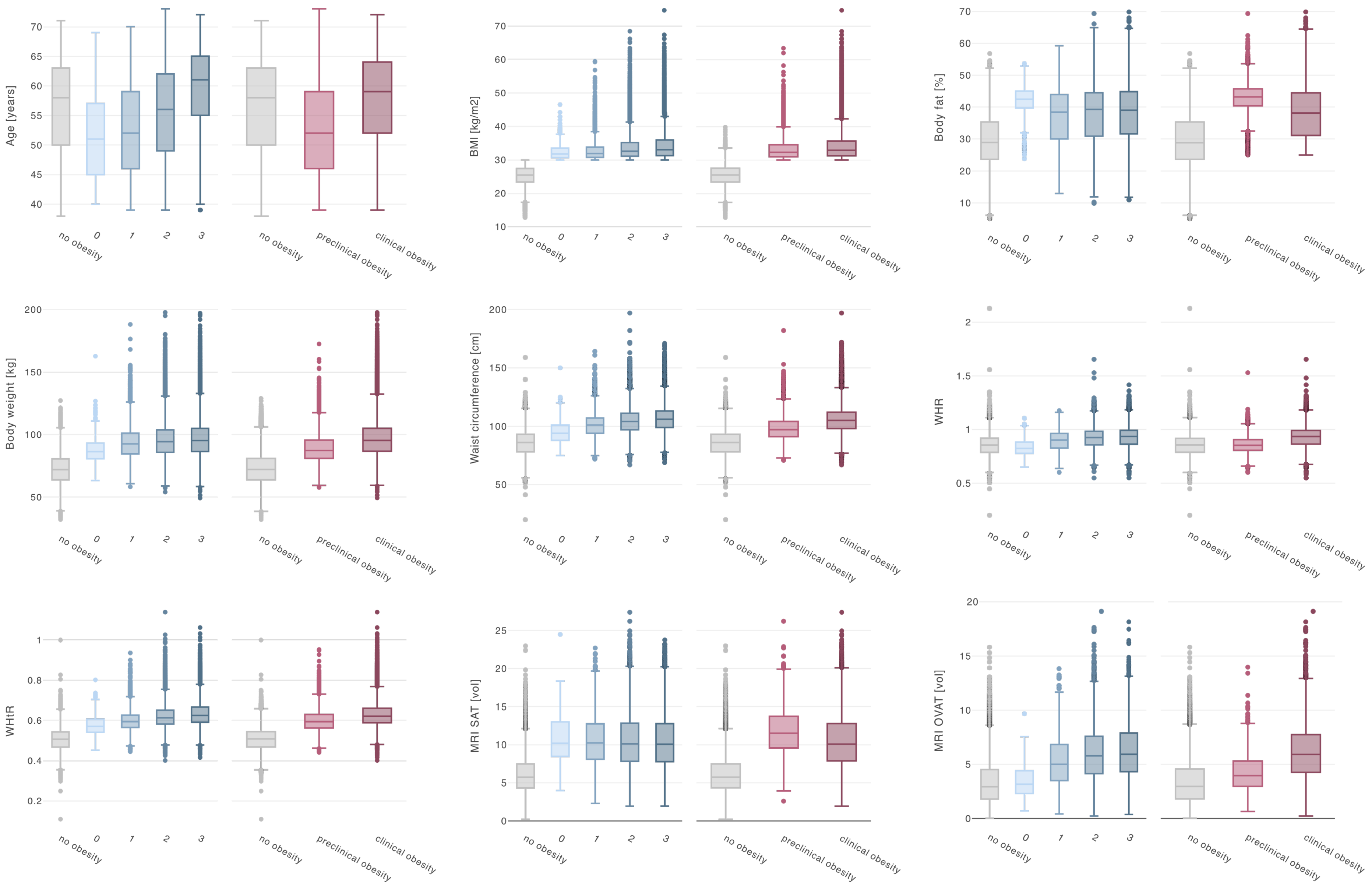

Age [years]
BMI [kg/m2]
Body fat [%]
Body weight [kg]
Waist circumference [cm]
WHR
WHtR
MRI SAT [vol]
MRI OVAT [vol]
no obesity
0
1
2
3
preclinical obesity
clinical obesity

**Suppl. Figure 2: Comparison of clinical parameters between the two obesity classification frameworks.** Box plots of selected clinical parameters displayed for the Edmonton Obesity Staging System (blue colors) and the Diagnostic Model for Obesity (red colors). The plots illustrate the distribution and differences in these variables both between the two assessment methods and within each individual grouping of the systems. Abbreviations: BMI: body mass index; MRI: magnetic resonance imaging; SAT: subcutaneous adipose tissue; VAT: visceral adipose tissue; WHtR: waist-to-heigh ratio; WHR: waist-to-hip ratio.
